# Temporal Trends in Artemisinin Partial Resistance and Other Antimalarial Drug Mutations in *Plasmodium falciparum* from Kagera Region, Northwestern Tanzania, 2021–2023

**DOI:** 10.1101/2025.11.26.25341086

**Authors:** Alfred Simkin, Salehe Mandai, Abebe A Fola, Neeva Wernsman Young, Dativa Pereus, Catherine Bakari, Rashid A Madebe, Misago D Seth, Rule B Mrengela, Angelina J Kisambale, Gervas A Chacha, Celine I Mandara, Filbert Francis, Daniel Mbwambo, Issa Garimo, Frank Chacky, Sijenunu Aaron, Abdallah Lusasi, Fabrizio Molteni, Ritha J A Njau, Stella Kajange, Samwel L Nhiga, Ally Mohamed, Jonathan J Juliano, Deus S Ishengoma, Jeffrey A Bailey

**Affiliations:** Department of Pathology and Laboratory Medicine, Brown University, Providence, RI, USA; Center for Computational Molecular Biology, Brown University, Providence, RI, USA; Ifakara Health Institute, Dar es Salaam, Tanzania; National Institute for Medical Research, Dar es Salaam, Tanzania; National Malaria Control Programme, Dodoma, Tanzania; Swiss Tropical Public Health Institute, Dar es Salaam, Tanzania; Department of Parasitology and Medical Entomology, Muhimbili University of Health and Allied Sciences, Dar es Salaam, Tanzania; President’s Office, Regional Administration and Local Government, Dodoma, Tanzania; Division of Infectious Diseases, University of North Carolina School of Medicine, University of North Carolina at Chapel Hill, Chapel Hill, NC, USA; Curriculum of Genetics and Molecular Biology, University of North Carolina School of Medicine, University of North Carolina at Chapel Hill, Chapel Hill, NC, USA; Institute for Global Health and Infectious Diseases, University of North Carolina School of Medicine, University of North Carolina at Chapel Hill, Chapel Hill, NC; Department of Epidemiology, Gillings School of Global Public Health, University of North Carolina at Chapel Hill, Chapel Hill, NC, USA

**Keywords:** malaria, *Plasmodium falciparum*, kelch, artemisinin, resistance, Tanzania, genomics, surveillance

## Abstract

Artemisinin-based combination therapies (ACTs) remain the cornerstone of malaria treatment, yet the emergence of artemisinin partial resistance (ART-R) in Africa threatens their efficacy. ART-R is primarily associated with mutations in the *Plasmodium falciparum kelch13* (K13) gene, notably R561**H**, which has been linked to delayed parasite clearance in East Africa. We conducted longitudinal molecular surveillance in Tanzania’s Kagera region from 2021 to 2023 to characterize temporal and spatial trends in ART-R and other antimalarial resistance markers. Using molecular inversion probes targeting key antimalarial resistance genes, we genotyped 2,826 isolates from seven districts. The WHO-validated K13 mutation R561**H** persisted in border districts of Karagwe and Kyerwa, with prevalence ranging from 14-26%, and appeared for the first time in Muleba (5.0%) and Bukoba rural district (0.7%) in 2023, indicating eastward spread toward Lake Victoria. Regional average prevalence of R561**H** rose from 5.5% in 2021 to 11.3% in 2022, then stabilized at 6.9% in 2023. Additional validated (A675**V**) and candidate (V568**G**, P441**L**) mutations were detected at low frequencies, suggesting ongoing diversification of the parasite population under local selection pressures. Partner-drug resistance markers showed minimal change: MDR1 **N**86Y remained near fixation, while CRT K76**T** declined from 5.9% (2021) to 2.4% (2023). Antifolate resistance was entrenched, with early DHFR and DHPS mutations near fixation and high-level resistance markers (DHFR I164**L** and DHPS A581**G**) exhibiting marked spatial heterogeneity, peaking at 38.1% and 48.1%, respectively, in eastern districts. These findings reveal micro-geographic heterogeneity in resistance and ongoing spread, emphasizing the need for district-level surveillance to detect emerging hotspots and guide interventions. Sustained molecular monitoring is critical to inform treatment policy, preserve ACT efficacy, and mitigate the risk of widespread resistance in East Africa.

Artemisinin-based combination therapies (ACTs) remain the cornerstone of malaria treatment, yet the emergence of artemisinin partial resistance (ART-R) in Africa threatens their efficacy. ART-R is primarily associated with mutations in the *Plasmodium falciparum* kelch13 (K13) gene, notably R561**H**, which has been linked to delayed parasite clearance in East Africa. We conducted longitudinal molecular surveillance in Tanzania’s Kagera region from 2021 to 2023 to characterize temporal and spatial trends in ART-R and other antimalarial resistance markers. Using molecular inversion probes targeting key antimalarial resistance genes, we genotyped 2,826 isolates from seven districts. The WHO-validated K13 mutation R561**H** persisted in border districts of Karagwe and Kyerwa, with prevalence ranging from 14-26%, and appeared for the first time in Muleba (5.0%) and Bukoba rural district (0.7%) in 2023, indicating eastward spread toward Lake Victoria. Regional average prevalence of R561**H** rose from 5.5% in 2021 to 11.3% in 2022, then stabilized at 6.9% in 2023. Additional validated (A675**V**) and candidate (V568**G**, P441**L**) mutations were detected at low frequencies, suggesting ongoing diversification of the parasite population under local selection pressures. Partner-drug resistance markers showed minimal change: MDR1 **N**86Y remained near fixation, while CRT K76**T** declined from 5.9% (2021) to 2.4% (2023). Antifolate resistance was entrenched, with early DHFR and DHPS mutations near fixation and high-level resistance markers (DHFR I164**L** and DHPS A581**G**) exhibiting marked spatial heterogeneity, peaking at 38.1% and 48.1%, respectively, in eastern districts. These findings reveal micro-geographic heterogeneity in resistance and ongoing spread, emphasizing the need for district-level surveillance to detect emerging hotspots and guide interventions. Sustained molecular monitoring is critical to inform treatment policy, preserve ACT efficacy, and mitigate the risk of widespread resistance in East Africa.

## INTRODUCTION

Africa is facing a looming public health crisis with the emergence of artemisinin partial resistance (ART-R). (Rosenthal et al., 2024) ART-R is associated with mutations in *Plasmodium falciparum kelch13* (K13), which have been linked to prolonged parasite clearance and longer clearance half-lives. Multiple World Health Organization (WHO) validated and candidate K13 polymorphisms, including P441**L**, C469**Y**/**F**, R561**H**, R622**I**, and A675**V**, are now detected widely in most countries along the Great Rift Valley.(Rosenthal et al., 2024) Another focus of P441**L** has also emerged in Southern Africa around the Namibia-Zambia border.(Eloff et al., 2025b; Martin et al., 2025) Mutations in partner drug resistance will be critical in clinical failure of artemisinin combination therapies (ACT), as was seen in Southeast Asia.(Björkman et al., 2024) As ACTs rely on artemisinin to reduce parasite burden, resistant parasites are increasingly exposed to partner drugs as monotherapy, heightening the risk of resistance development.(Rosenthal et al., 2024) Additionally, existing partner drug resistance mutations in Africa could further accelerate the spread of ART-R, mirroring the pattern observed with mefloquine resistance in Asia.(Björkman et al., 2024) Understanding the spread of ART-R and partner drug mutations requires samples collected across multiple years in affected areas. This information is essential for malaria control planning in order to implement potential interventions that may reduce resistance, such as multiple first-line therapy, triple or sequential ACT, non-ACT antimalarial drugs, or potentially malaria vaccination.

The Kagera Region in Northwest Tanzania appears to be part of the outward spread of R561**H** K13 mutation from a likely origin in Rwanda. After initial detection of the polymorphism in Rwanda in samples collected in 2014, a therapeutic efficacy study (TES) in 2018 confirmed clinical ART-R.(Uwimana et al., 2020, 2021) Subsequent analysis of samples collected from asymptomatic individuals in the 2014 Rwanda Demographic Health Survey showed the mutation occurring along the border with Tanzania.(Kirby et al., 2023) In 2021, the R561**H** mutation was found in multiple clinics in Kagera along the Rwanda border, with a prevalence of 22.8% [31 of 136] in Karagwe, 14.4% [17 of 118]) in Kyerwa, and 1.4% [two of 144] in Ngara.(Juliano et al., 2024) A TES in the same region in 2022 documented clinical ART-R, based on delayed clearance with the median parasite clearance half-life in patients harbouring parasites with R561**H** mutation reaching over 6 hours in all treatment groups, further confirming its clinical impact.(Ishengoma et al., 2024)

Beyond ART-R, the efficacy of other antimalarials is at risk. Non-synonymous polymorphisms in *P. falciparum* multidrug resistance (MDR1) protein and *P. falciparum* chloroquine resistance transporter (CRT) have been linked to reduced susceptibility to amodiaquine (Humphreys et al., 2007) and lumefantrine (Venkatesan et al., 2014). Markers for lumefantrine resistance remain unclear (Rosado et al., 2024), but the wild type MDR1 **N**86Y allele has been associated with re-infection following artemether-lumefantrine (AL) treatment.(Sisowath et al., 2005) As lumefantrine and amodiaquine are the most widely used ACT partner drugs across Africa, monitoring genetic mutations linked to reduced susceptibility is crucial. Mutations in *P. falciparum* dihydrofolate reductase (DHFR) and *P. falciparum* dihydropteroate synthase (DHPS) impact susceptibility to pyrimethamine (P) and sulfadoxine (S), respectively. Mutations conferring high-level resistance to SP, a combination used frequently in chemoprevention, are also important to monitor, in particular DHFR N51**I**, C59**R**, S108**N/T**, and I164**L**, which together form the DHFR **IRNL** resistance haplotype, and DHPS A437**G**, DHPS K540**E**, and DHPS A581**G**, which together form the DHPS **GEG** resistance haplotype. In each of these genes, these haplotypes have formed through sequential mutations, with the latest mutations of DHFR I164**L** and DHPS A581**G** demonstrating the greatest pyrimethamine and sulfadoxine resistance, respectively.(Hastings et al., 2002; Mousa et al., 2025)

To understand the dynamics of artemisinin partial resistance and associated partner drug mutations in this emerging hotspot, we conducted annual cross-sectional surveys within the ongoing malaria molecular surveillance in Tanzania’s Kagera region from 2021 to 2023. By analyzing temporal and spatial trends in validated and candidate kelch13 polymorphisms alongside markers for partner drug and antifolate resistance and tolerance [DHFR I164**L**, DHPS A581**G**, CRT K76**T**, and MDR1 **N**86Y], this study provides critical insights into the evolution and spread of antimalarial drug resistance in the region. These findings are essential for informing malaria control strategies, guiding drug policy decisions, and supporting interventions—such as multiple first-line therapies, triple ACT regimens, future deployment of non-ACT antimalarial drugs, or integration with malaria vaccination—to preserve the efficacy of current treatments and mitigate the threat of widespread resistance.

## METHODS

### Ethics Statement

This study was conducted as part of the Molecular Surveillance of Malaria in Tanzania (MSMT) project, whose protocol was submitted to, reviewed, and approved by the Medical Research Coordinating Committee (MRCC) of the National Institute for Medical Research (NIMR) in Tanzania (NIMR/HQ/R.8a/Vol.IX/3579). All research participants provided individual consent (or assent for children aged 7–17 years) for both participation in the survey and biobanking for future research. For participants under the legal age of adulthood in Tanzania (<18 years), consent was obtained from a parent or guardian. The informed consent form was developed in English, translated into Kiswahili, and used to obtain consent verbally and in writing. Participants either signed the consent or assent form or, if illiterate, provided a thumbprint accompanied by the signature of an independent witness.

### Patient samples

The MSMT project collected capillary dried blood spot (DBS) samples from patients aged six months and older who tested positive for malaria via rapid diagnostic tests at participating clinics. The 2021 and 2022 surveys covered 100 health facilities across ten regions: Dar es Salaam, Dodoma, Kagera, Kilimanjaro, Manyara, Mara, Mtwara, Njombe, Songwe, and Tabora while the 2023 survey covered all 26 regions of Mainland Tanzania; here, we focus only on clinics in the Kagera region. The sites in Kagera were selected to be able to evaluate the border region with Rwanda, where K13 polymorphisms were previously described, as well as to assess if spread was occurring to the East toward Lake Victoria and South toward Lake Tanganyika. Samples were collected between 01 February and 26 July, 01 February and 03 August, and 23 January and 28 August in 2021, 2022, and 2023, respectively. In 2021 and 2023, one site in each district was sampled in Bukoba, Biharamulo, Karagwe, and Misenyi districts, while two sites were covered in Muleba, Kyerwa, and Ngara. In 2022, one site in each of 3 districts was sampled (Muleba, Karagwe, and Ngara). Written informed consent was obtained as approved by the Tanzanian Medical Research Coordinating Committee of NIMR. Deidentified dried blood spot (DBS) samples from 2021 were processed at Brown University, and 2022 and 2023 were processed at the National Institute for Medical Research (NIMR) in Dar es Salaam, Tanzania.

### DNA Extraction and Sequencing

DNA was extracted using the Chelex-Tween protocol, followed by molecular inversion probe (MIP) capture. The resulting library was purified using the New England Biolabs (NEB) Gel extraction kit (NEB Inc., Ipswich, MA, USA). The NEBNext® Library Quant Kit for Illumina® (NEB Inc., Ipswich, MA, USA) was used for qPCR-based library quantitation and sequenced using Illumina platforms, MiSeq & NextSeq (Illumina, Inc., San Diego, CA, USA), using 3D7 and 7G8 as controls (MRA-102G and 152G, BEI Resources, Manassas, VA). The sequencing reads were then transferred to the main server for downstream bioinformatics analysis. Libraries lacking sufficient read depth were rebalanced and resequenced.

### Parasite Genotyping

We attempted sequencing of 575 isolates from 3 districts in 2022 and 1687 isolates from 7 districts in 2023. Samples from 2021 have been previously reported.(Juliano et al., 2024) Samples from 2022 and 2023 were genotyped with a smaller panel containing a subset of 121 and 184 MIP probes, targeting major antimalarial resistance polymorphisms, respectively. For analysis, we limited data to the 121 targets from the smallest MIP panel.

### Bioinformatics Analysis

Variant calling was performed using MIPTools to process fastq data, with variants identified through FreeBayes as described previously (Juliano et al., 2024). Downstream filtering leveraged unique molecular index (UMI) clustering for error correction, ensuring high-confidence calls and quantification. Antimalarial resistance allele prevalence for known mutations was calculated for sites with a MIP UMI count of three or more. Heterozygous variants required an alternate allele count of at least one UMI. For other emerging novel mutations, a MIP UMI count of 10 with an alternate allele count of 3 was required.

### Data Analysis

Descriptive statistics were used to summarize sample characteristics, including frequencies and proportions of successfully genotyped isolates across years and districts. Prevalence estimates and 95% confidence intervals (CIs) were calculated for key mutations associated with antimalarial drug resistance, stratified by year and district. Temporal changes in mutation frequencies were evaluated using tests for proportions and illustrated with trend plots. Spatial patterns of resistance mutations were analyzed by the district to detect geographic emergence and expansion. All data visualizations, including maps and mutation trend plots, were generated using the ggplot2 and sf R packages; map shapefiles were obtained from GADM.org. Statistical analyses were conducted using R version 4.2.1 (The R Foundation for Statistical Computing, Vienna, Austria). A P-value < 0.05 was considered statistically significant.

For analysis, we leveraged previously reported genotyping data from 942 successfully genotyped samples from Kagera from 2021. Prevalences were calculated at the district level. Because genotyping success varied by locus, unweighted prevalence estimates were calculated using locus-specific denominators (*p*lJ=lJ*x*/*n*lJ×lJ100, where *p* is the prevalence, *x* is the number of samples with mutant alleles and *n* is the number of successfully genotyped samples). Unweighted prevalence was calculated using the miplicorn R package v.0.2.90 (https://github.com/bailey-lab/miplicorn). Maps were generated using the sf package in R 4.2.1.

## RESULTS

### Characteristics of successfully genotyped samples

Overall, 2,979 Kagera samples were collected and sequenced over three years. Of the attempted samples, 2,826 (94.8%) isolates passed filters for MIP extraction. Overall, 534 isolates from 2022 and 1,374 isolates from 2023 passed filtering and were compared to 918 previously published isolates from 2021 (Juliano et al., 2024). Sampling in Kagera was at 11 sites representing 7 districts, with interim sampling in 2022 of only 3 districts (**Figure 1**).

**Figure 1.**
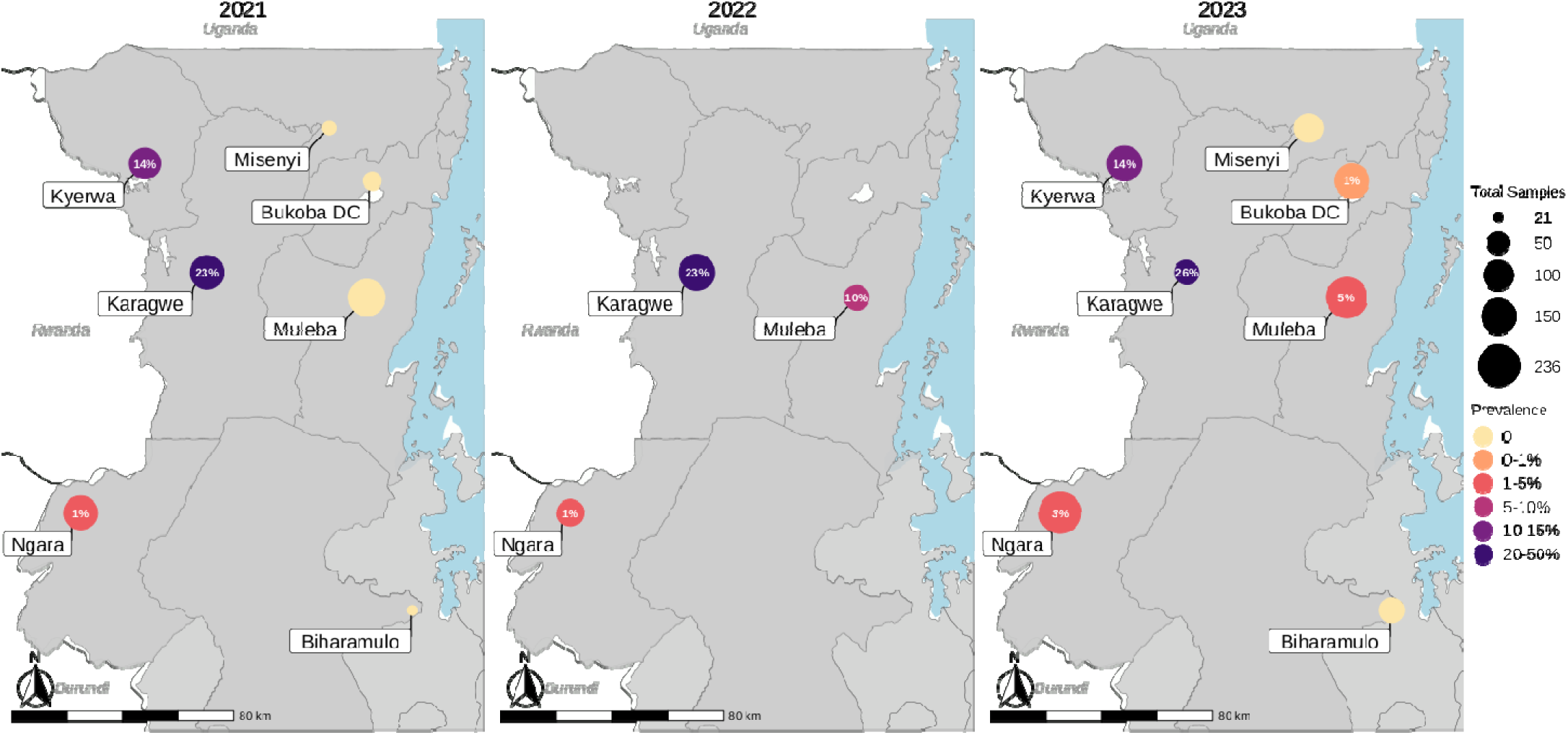
Spatial Distribution and Temporal Changes in K13 R561H. Prevalences are shown as percentages (inside of circles) and as a color scale from light (low prevalence) to dark (high prevalence), while sample counts are represented by point size. Data is aggregated by District.

### ART-R K13 polymorphisms

K13 R561**H** mutations were commonly found from 2021-2023, with an unweighted average district prevalence across the Kagera region rising over two years from 5.5% in 2021 to 6.9% in 2023 (**Table 1).** Examining the spatiotemporal relationships, Karagwe and Ngara, along the Rwanda border, showed slight increases in prevalence for K13 R561**H** over this time (**Figure 1**). The most notable differences in prevalence occurred in two districts in the East, where R561**H** mutations were detected for the first time in 2023, consistent with further spread. The prevalence of K13 R561**H** in Muleba district went from 0% (0/171) to 5% (11/220, CI 2.5%-8.8%), and Bukoba district went from 0% (0/35) to 0.7% (1/147, CI 0.0%-3.7%). The only other validated ART-R K13 mutations to be consistently found across all 3 years were mutants A675**V** and V568**G** (**Table 1**). The A675**V** isolates were all in Karagwe (all years), as well as Muleba and Kyerwa in 2023. The V568**G** isolates were found in Bukoba in 2021 and 2023, and also in Muleba in 2021 and 2022, and in Kyerwa in 2021. R622**I**, known to be at high prevalence in Eritrea and Ethiopia (Fola et al., 2023; Mihreteab et al., 2023), was found at very low levels in Misenyi in 2023. The mutation P441**L**, found commonly in Zambia, Namibia, Ethiopia, Rwanda and Uganda (Conrad et al., 2023; Eloff et al., 2025a; Martin et al., 2025; Wernsman Young et al., 2025), was found in 2021 and 2023 in Kyerwa, and in 2023 in Misenyi (**Table 1**). In addition, we have coverage information for nine other validated or candidate WHO ART-R K13 mutations that were assayed and found to be absent from Kagera (**Table S1**). Several non-validated non-synonymous K13 mutations were also identified (**Table S2; Figure S1**), with more of these mutations occurring in 2022 and 2023 than in 2021. These mutations were found within Bukoba, Karagwe, Kyerwa, Misenyi, and Ngara, with especially elevated numbers of mutated samples in Karagwe in 2021 and in Ngara in 2023 relative to background sampling rates (binomial P-values of 1.2*10^-4^ and 6.1*10^-5^, respectively).

**Table 1.** District Level Prevalence of Validated and Candidate K13 Polymorphisms 2021-2023.

|  |  | Biharamulo |  | Bukoba DC |  | Karagwe |  | Kyerwa |  | Misenyi |  | Muleba |  | Ngara |  | District Average* |
| --- | --- | --- | --- | --- | --- | --- | --- | --- | --- | --- | --- | --- | --- | --- | --- | --- |
| mutation | year | freq | prev (CI) | freq | prev (CI) | freq | prev (CI) | freq | prev (CI) | freq | prev (CI) | freq | prev (CI) | freq | prev (CI) | prev |
| k13 P441L | 2021 | 0/21 | 0.0 (0.0-16.1) | 0/38 | 0.0 (0.0-9.3) | 0/138 | 0.0 (0.0-2.6) | 1/120 | 0.8 (0.0-4.6) | 0/26 | 0.0 (0.0-13.2) | 0/173 | 0.0 (0.0-2.1) | 0/155 | 0.0 (0.0-2.4) | 0.1 |
|  | 2022 | - | - | - | - | 0/157 | 0.0 (0.0-2.3) | - | - | - | - | 0/67 | 0.0 (0.0-5.4) | 0/90 | 0.0 (0.0-4.0) | 0 |
|  | 2023 | 0/65 | 0.0 (0.0-5.5) | 0/146 | 0.0 (0.0-2.5) | 0/63 | 0.0 (0.0-5.7) | 2/155 | 1.3 (0.2-4.6) | 3/95 | 3.2 (0.7-9.0) | 0/202 | 0.0 (0.0-1.8) | 0/223 | 0.0 (0.0-1.6) | 0.6 |
| k13 F446I | 2021 | 0/21 | 0.0 (0.0-16.1) | 0/38 | 0.0 (0.0-9.3) | 0/138 | 0.0 (0.0-2.6) | 0/120 | 0.0 (0.0-3.0) | 1/26 | 3.8 (0.1-19.6) | 0/173 | 0.0 (0.0-2.1) | 0/155 | 0.0 (0.0-2.4) | 0.5 |
|  | 2022 | - | - | - | - | 0/157 | 0.0 (0.0-2.3) | - | - | - | - | 0/67 | 0.0 (0.0-5.4) | 0/90 | 0.0 (0.0-4.0) | 0 |
|  | 2023 | 0/65 | 0.0 (0.0-5.5) | 0/146 | 0.0 (0.0-2.5) | 0/63 | 0.0 (0.0-5.7) | 0/155 | 0.0 (0.0-2.4) | 0/95 | 0.0 (0.0-3.8) | 0/202 | 0.0 (0.0-1.8) | 0/223 | 0.0 (0.0-1.6) | 0 |
| k13 G449A | 2021 | 0/21 | 0.0 (0.0-16.1) | 0/38 | 0.0 (0.0-9.3) | 0/138 | 0.0 (0.0-2.6) | 0/120 | 0.0 (0.0-3.0) | 0/26 | 0.0 (0.0-13.2) | 0/173 | 0.0 (0.0-2.1) | 0/155 | 0.0 (0.0-2.4) | 0 |
|  | 2022 | - | - | - | - | 0/157 | 0.0 (0.0-2.3) | - | - | - | - | 0/67 | 0.0 (0.0-5.4) | 0/90 | 0.0 (0.0-4.0) | 0 |
|  | 2023 | 0/65 | 0.0 (0.0-5.5) | 0/146 | 0.0 (0.0-2.5) | 0/63 | 0.0 (0.0-5.7) | 6/155 | 3.9 (1.4-8.2) | 0/95 | 0.0 (0.0-3.8) | 0/202 | 0.0 (0.0-1.8) | 0/223 | 0.0 (0.0-1.6) | 0.6 |
| k13 C469Y | 2021 | 0/21 | 0.0 (0.0-16.1) | 0/38 | 0.0 (0.0-9.3) | 0/138 | 0.0 (0.0-2.6) | 0/120 | 0.0 (0.0-3.0) | 0/26 | 0.0 (0.0-13.2) | 0/173 | 0.0 (0.0-2.1) | 0/155 | 0.0 (0.0-2.4) | 0 |
|  | 2022 | - | - | - | - | 0/157 | 0.0 (0.0-2.3) | - | - | - | - | 0/67 | 0.0 (0.0-5.4) | 0/90 | 0.0 (0.0-4.0) | 0 |
|  | 2023 | 0/65 | 0.0 (0.0-5.5) | 0/146 | 0.0 (0.0-2.5) | 1/63 | 1.6 (0.0-8.5) | 0/155 | 0.0 (0.0-2.4) | 0/95 | 0.0 (0.0-3.8) | 0/202 | 0.0 (0.0-1.8) | 0/223 | 0.0 (0.0-1.6) | 0.2 |
| k13 P527H | 2021 | 0/21 | 0.0 (0.0-16.1) | 1/39 | 2.6 (0.1-13.5) | 0/146 | 0.0 (0.0-2.5) | 0/132 | 0.0 (0.0-2.8) | 0/24 | 0.0 (0.0-14.2) | 0/184 | 0.0 (0.0-2.0) | 0/174 | 0.0 (0.0-2.1) | 0.4 |
|  | 2022 | - | - | - | - | 0/170 | 0.0 (0.0-2.1) | - | - | - | - | 0/82 | 0.0 (0.0-4.4) | 1/96 | 1.0 (0.0-5.7) | 0.3 |
|  | 2023 | 0/74 | 0.0 (0.0-4.9) | 0/155 | 0.0 (0.0-2.4) | 0/66 | 0.0 (0.0-5.4) | 0/170 | 0.0 (0.0-2.1) | 0/104 | 0.0 (0.0-3.5) | 0/229 | 0.0 (0.0-1.6) | 0/251 | 0.0 (0.0-1.5) | 0 |
| k13 N537D | 2021 | 0/21 | 0.0 (0.0-16.1) | 0/39 | 0.0 (0.0-9.0) | 0/146 | 0.0 (0.0-2.5) | 0/132 | 0.0 (0.0-2.8) | 0/24 | 0.0 (0.0-14.2) | 0/184 | 0.0 (0.0-2.0) | 0/174 | 0.0 (0.0-2.1) | 0 |
|  | 2022 | - | - | - | - | 0/170 | 0.0 (0.0-2.1) | - | - | - | - | 0/82 | 0.0 (0.0-4.4) | 0/96 | 0.0 (0.0-3.8) | 0 |
|  | 2023 | 0/74 | 0.0 (0.0-4.9) | 0/155 | 0.0 (0.0-2.4) | 0/66 | 0.0 (0.0-5.4) | 1/170 | 0.6 (0.0-3.2) | 0/104 | 0.0 (0.0-3.5) | 0/229 | 0.0 (0.0-1.6) | 0/251 | 0.0 (0.0-1.5) | 0.1 |
| k13 N537I | 2021 | 0/21 | 0.0 (0.0-16.1) | 0/39 | 0.0 (0.0-9.0) | 0/146 | 0.0 (0.0-2.5) | 0/132 | 0.0 (0.0-2.8) | 0/24 | 0.0 (0.0-14.2) | 0/184 | 0.0 (0.0-2.0) | 0/174 | 0.0 (0.0-2.1) | 0 |
|  | 2022 | - | - | - | - | 0/170 | 0.0 (0.0-2.1) | - | - | - | - | 0/82 | 0.0 (0.0-4.4) | 0/96 | 0.0 (0.0-3.8) | 0 |
|  | 2023 | 0/74 | 0.0 (0.0-4.9) | 0/155 | 0.0 (0.0-2.4) | 0/66 | 0.0 (0.0-5.4) | 0/170 | 0.0 (0.0-2.1) | 0/104 | 0.0 (0.0-3.5) | 1/229 | 0.4 (0.0-2.4) | 0/251 | 0.0 (0.0-1.5) | 0.1 |
| k13 G538V | 2021 | 0/21 | 0.0 (0.0-16.1) | 0/39 | 0.0 (0.0-9.0) | 0/146 | 0.0 (0.0-2.5) | 0/132 | 0.0 (0.0-2.8) | 0/24 | 0.0 (0.0-14.2) | 0/184 | 0.0 (0.0-2.0) | 0/174 | 0.0 (0.0-2.1) | 0 |
|  | 2022 | - | - | - | - | 0/170 | 0.0 (0.0-2.1) | - | - | - | - | 0/82 | 0.0 (0.0-4.4) | 0/96 | 0.0 (0.0-3.8) | 0 |
|  | 2023 | 0/74 | 0.0 (0.0-4.9) | 0/155 | 0.0 (0.0-2.4) | 0/66 | 0.0 (0.0-5.4) | 0/170 | 0.0 (0.0-2.1) | 0/104 | 0.0 (0.0-3.5) | 1/229 | 0.4 (0.0-2.4) | 0/251 | 0.0 (0.0-1.5) | 0.1 |
| k13 I543T | 2021 | 0/21 | 0.0 (0.0-16.1) | 0/39 | 0.0 (0.0-9.0) | 0/146 | 0.0 (0.0-2.5) | 0/132 | 0.0 (0.0-2.8) | 0/24 | 0.0 (0.0-14.2) | 0/184 | 0.0 (0.0-2.0) | 0/174 | 0.0 (0.0-2.1) | 0 |
|  | 2022 | - | - | - | - | 0/170 | 0.0 (0.0-2.1) | - | - | - | - | 0/82 | 0.0 (0.0-4.4) | 0/96 | 0.0 (0.0-3.8) | 0 |
|  | 2023 | 0/74 | 0.0 (0.0-4.9) | 1/155 | 0.6 (0.0-3.5) | 0/66 | 0.0 (0.0-5.4) | 1/170 | 0.6 (0.0-3.2) | 0/104 | 0.0 (0.0-3.5) | 0/229 | 0.0 (0.0-1.6) | 0/251 | 0.0 (0.0-1.5) | 0.2 |
| k13 R561H | 2021 | 0/21 | 0.0 (0.0-16.1) | 0/35 | 0.0 (0.0-10.0) | 31/136 | 22.8 (16.0-30.8) | 17/118 | 14.4 (8.6-22.1) | 0/24 | 0.0 (0.0-14.2) | 0/171 | 0.0 (0.0-2.1) | 2/144 | 1.4 (0.2-4.9) | 5.5 |
|  | 2022 | - | - | - | - | 37/162 | 22.8 (16.6-30.1) | - | - | - | - | 7/70 | 10.0 (4.1-19.5) | 1/83 | 1.2 (0.0-6.5) | 11.3 |
|  | 2023 | 0/70 | 0.0 (0.0-5.1) | 1/147 | 0.7 (0.0-3.7) | 16/62 | 25.8 (15.5-38.5) | 22/158 | 13.9 (8.9-20.3) | 0/98 | 0.0 (0.0-3.7) | 11/220 | 5.0 (2.5-8.8) | 7/236 | 3.0 (1.2-6.0) | 6.9 |
| k13 V568G | 2021 | 0/21 | 0.0 (0.0-16.1) | 2/35 | 5.7 (0.7-19.2) | 0/136 | 0.0 (0.0-2.7) | 1/118 | 0.8 (0.0-4.6) | 0/24 | 0.0 (0.0-14.2) | 0/171 | 0.0 (0.0-2.1) | 1/144 | 0.7 (0.0-3.8) | 1 |
|  | 2022 | - | - | - | - | 0/162 | 0.0 (0.0-2.3) | - | - | - | - | 0/70 | 0.0 (0.0-5.1) | 1/83 | 1.2 (0.0-6.5) | 0.4 |
|  | 2023 | 0/70 | 0.0 (0.0-5.1) | 1/147 | 0.7 (0.0-3.7) | 0/62 | 0.0 (0.0-5.8) | 0/158 | 0.0 (0.0-2.3) | 0/98 | 0.0 (0.0-3.7) | 0/220 | 0.0 (0.0-1.7) | 0/236 | 0.0 (0.0-1.6) | 0.1 |
| k13 P574L | 2021 | 0/21 | 0.0 (0.0-16.1) | 0/35 | 0.0 (0.0-10.0) | 0/136 | 0.0 (0.0-2.7) | 0/118 | 0.0 (0.0-3.1) | 0/24 | 0.0 (0.0-14.2) | 0/171 | 0.0 (0.0-2.1) | 0/144 | 0.0 (0.0-2.5) | 0 |
|  | 2022 | - | - | - | - | 0/162 | 0.0 (0.0-2.3) | - | - | - | - | 0/70 | 0.0 (0.0-5.1) | 1/83 | 1.2 (0.0-6.5) | 0.4 |
|  | 2023 | 0/70 | 0.0 (0.0-5.1) | 0/147 | 0.0 (0.0-2.5) | 0/62 | 0.0 (0.0-5.8) | 0/158 | 0.0 (0.0-2.3) | 0/98 | 0.0 (0.0-3.7) | 1/220 | 0.5 (0.0-2.5) | 1/236 | 0.4 (0.0-2.3) | 0.1 |
| k13 R622I | 2021 | 0/21 | 0.0 (0.0-16.1) | 0/39 | 0.0 (0.0-9.0) | 0/148 | 0.0 (0.0-2.5) | 0/127 | 0.0 (0.0-2.9) | 0/27 | 0.0 (0.0-12.8) | 0/183 | 0.0 (0.0-2.0) | 0/168 | 0.0 (0.0-2.2) | 0 |
|  | 2022 | - | - | - | - | 0/166 | 0.0 (0.0-2.2) | - | - | - | - | 0/84 | 0.0 (0.0-4.3) | 0/99 | 0.0 (0.0-3.7) | 0 |
|  | 2023 | 0/70 | 0.0 (0.0-5.1) | 0/153 | 0.0 (0.0-2.4) | 0/65 | 0.0 (0.0-5.5) | 0/162 | 0.0 (0.0-2.3) | 1/101 | 1.0 (0.0-5.4) | 0/216 | 0.0 (0.0-1.7) | 0/242 | 0.0 (0.0-1.5) | 0.1 |
| k13 A675V | 2021 | 0/21 | 0.0 (0.0-16.1) | 0/41 | 0.0 (0.0-8.6) | 1/138 | 0.7 (0.0-4.0) | 0/122 | 0.0 (0.0-3.0) | 0/26 | 0.0 (0.0-13.2) | 0/173 | 0.0 (0.0-2.1) | 0/162 | 0.0 (0.0-2.3) | 0.1 |
|  | 2022 | - | - | - | - | 1/170 | 0.6 (0.0-3.2) | - | - | - | - | 0/86 | 0.0 (0.0-4.2) | 0/94 | 0.0 (0.0-3.8) | 0.2 |
|  | 2023 | 0/66 | 0.0 (0.0-5.4) | 0/143 | 0.0 (0.0-2.5) | 3/62 | 4.8 (1.0-13.5) | 1/153 | 0.7 (0.0-3.6) | 0/97 | 0.0 (0.0-3.7) | 1/199 | 0.5 (0.0-2.8) | 0/224 | 0.0 (0.0-1.6) | 0.9 |

#: The data presented is the number of samples containing mutant alleles over the total number successfully genotyped (obs/pop), the calculated prevalence, expressed as a percentage (prev), and the 95% confidence interval for the prevalence as determined by the exact (also known as beta) approximation interval method, as implemented by the Python statsmodels module (v.0.14.4), also expressed as a percentage (CI). \*District Average is the equally weighted average of the districts for the region.

### Changes in mutations associated with tolerance to partner drugs and anti-folates

Overall, the changes in partner drug mutations were small. The MDR1 **N**86Y wild type allele (N), associated with tolerance to lumefantrine (Malmberg et al., 2013), is near fixation in all districts across the years (**Table 2**). Overall, the K76**T** mutant allele of CRT, associated with amodiaquine resistance (Olliaro et al., 1996), occurred at a low frequency in the population, with a decrease from 5.9% in 2021 to 2.4% in 2023 (**Table 2**). This was primarily driven by a decrease in Ngara, which decreased from 24.2% (CI: 17.1%-32.6%) to 9.7% (CI: 6.1%-14.5%) over this period (**Figure 2C**).

**Figure 2.**
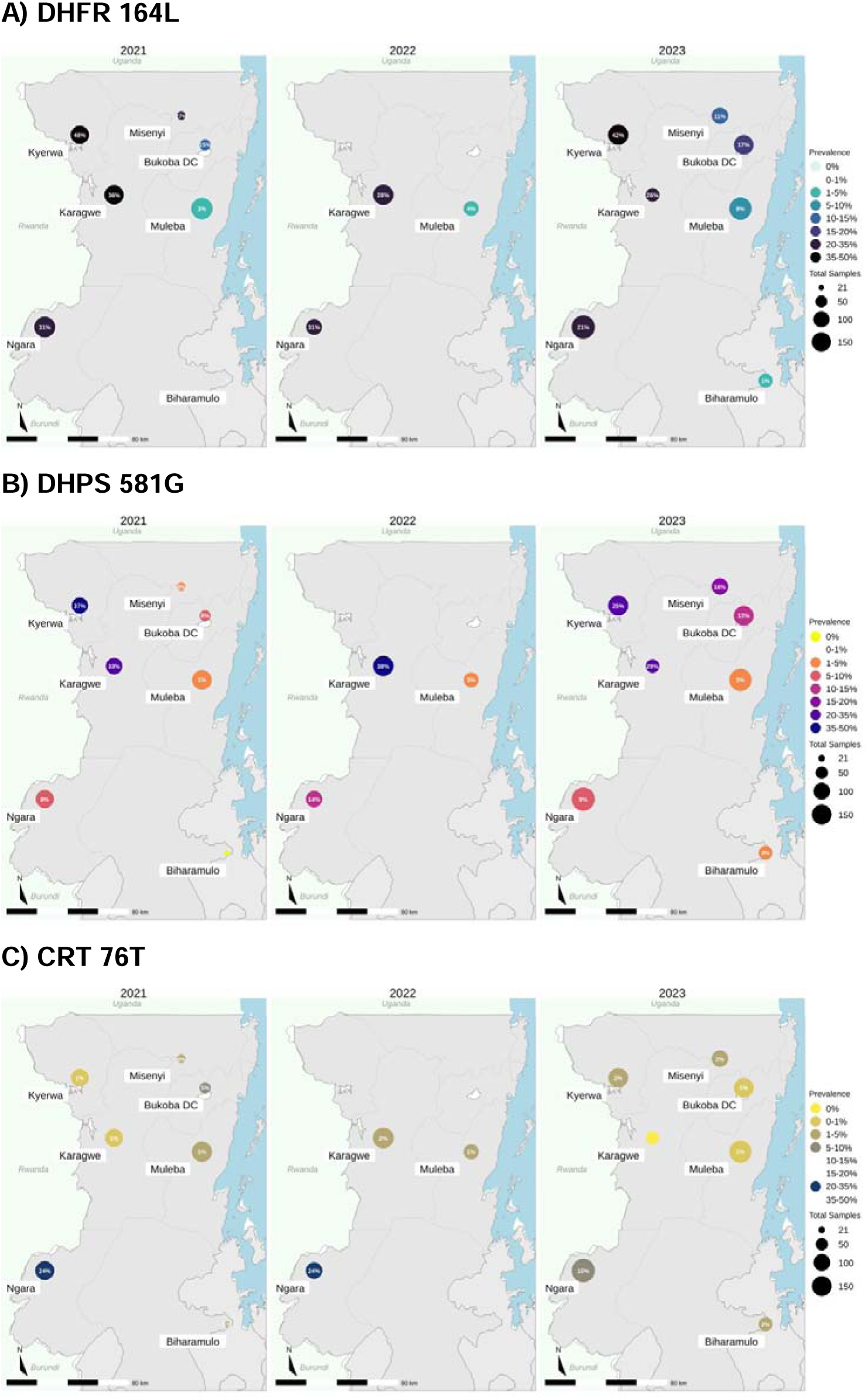
Key Mutations for sulfadoxine-pyremathimine and chloroquine resistance and their spatio-temporal correlation. The pyrimethamine resistance mutation DHFR 164L (**Panel A**), sulfadoxine resistance mutation DHPS 581G (**Panel B**), and chloroquine resistance CRT 76T (**Panel C**) are shown across years and sites. Prevalences are shown as percentages (in parentheses) along with the underlying sample counts.

**Table 2.** District level prevalence of partner drug (MDR1 and CRT), DHFR and DHPS mutations of interest.

|  |  | Biharamulo |  | Bukoba DC |  | Karagwe |  | Kyerwa |  | Misenyi |  | Muleba |  | Ngara |  | District Average* |
| --- | --- | --- | --- | --- | --- | --- | --- | --- | --- | --- | --- | --- | --- | --- | --- | --- |
| mutation | year | freq | prev (CI) | freq | prev (CI) | freq | prev (CI) | freq | prev (CI) | freq | prev (CI) | freq | prev (CI) | freq | prev (CI) | prev |
| crt<br>K76T | 2021 | 1/20 | 5.0 (0.1-24.9) | 2/38 | 5.3 (0.6-17.7) | 1/116 | 0.9 (0.0-4.7) | 1/110 | 0.9 (0.0-5.0) | 1/25 | 4.0 (0.1-20.4) | 2/152 | 1.3 (0.2-4.7) | 31/128 | 24.2 (17.1-32.6) | 5.9 |
|  | 2022 | - | - | - | - | 3/164 | 1.8 (0.4-5.3) | - | - | - | - | 1/76 | 1.3 (0.0-7.1) | 23/94 | 24.5 (16.2-34.4) | 9.2 |
|  | 2023 | 1/61 | 1.6 (0.0-8.8) | 1/144 | 0.7 (0.0-3.8) | 0/56 | 0.0 (0.0-6.4) | 3/144 | 2.1 (0.4-6.0) | 2/94 | 2.1 (0.3-7.5) | 1/190 | 0.5 (0.0-2.9) | 21/216 | 9.7 (6.1-14.5) | 2.4 |
| dhfr<br>I164L | 2021 | 0/20 | 0.0 (0.0-16.8) | 3/37 | 8.1 (1.7-21.9) | 31/95 | 32.6 (23.4-43.0) | 33/89 | 37.1 (27.1-48.0) | 1/25 | 4.0 (0.1-20.4) | 2/142 | 1.4 (0.2-5.0) | 10/118 | 8.5 (4.1-15.0) | 13.1 |
|  | 2022 | - | - | - | - | 59/155 | 38.1 (30.4-46.2) | - | - | - | - | 2/73 | 2.7 (0.3-9.5) | 13/94 | 13.8 (7.6-22.5) | 18.2 |
|  | 2023 | 1/62 | 1.6 (0.0-8.7) | 19/147 | 12.9 (8.0-19.4) | 17/58 | 29.3 (18.1-42.7) | 37/150 | 24.7 (18.0-32.4) | 15/95 | 15.8 (9.1-24.7) | 5/194 | 2.6 (0.8-5.9) | 20/219 | 9.1 (5.7-13.8) | 13.7 |
| dhps<br>A581G | 2021 | 0/21 | 0.0 (0.0-16.1) | 6/40 | 15.0 (5.7-29.8) | 54/150 | 36.0 (28.3-44.2) | 62/129 | 48.1 (39.2-57.0) | 7/26 | 26.9 (11.6-47.8) | 6/190 | 3.2 (1.2-6.7) | 55/177 | 31.1 (24.3-38.5) | 22.9 |
|  | 2022 | - | - | - | - | 47/168 | 28.0 (21.3-35.4) | - | - | - | - | 3/81 | 3.7 (0.8-10.4) | 28/91 | 30.8 (21.5-41.3) | 20.8 |
|  | 2023 | 1/70 | 1.4 (0.0-7.7) | 26/153 | 17.0 (11.4-23.9) | 17/65 | 26.2 (16.0-38.5) | 69/166 | 41.6 (34.0-49.5) | 11/100 | 11.0 (5.6-18.8) | 18/209 | 8.6 (5.2-13.3) | 49/235 | 20.9 (15.8-26.6) | 18.1 |
| mdr1<br>N86Y | 2021 | 0/20 | 0.0 (0.0-16.8) | 0/38 | 0.0 (0.0-9.3) | 0/148 | 0.0 (0.0-2.5) | 1/121 | 0.8 (0.0-4.5) | 0/26 | 0.0 (0.0-13.2) | 0/180 | 0.0 (0.0-2.0) | 2/179 | 1.1 (0.1-4.0) | 0.3 |
|  | 2022 | - | - | - | - | 0/155 | 0.0 (0.0-2.4) | - | - | - | - | 0/64 | 0.0 (0.0-5.6) | 0/79 | 0.0 (0.0-4.6) | 0 |
|  | 2023 | 0/65 | 0.0 (0.0-5.5) | 1/141 | 0.7 (0.0-3.9) | 0/60 | 0.0 (0.0-6.0) | 0/150 | 0.0 (0.0-2.4) | 0/96 | 0.0 (0.0-3.8) | 0/191 | 0.0 (0.0-1.9) | 1/215 | 0.5 (0.0-2.6) | 0.2 |

#: The data presented is the number of samples containing mutant alleles over the total number successfully genotyped (obs/pop), the calculated prevalence, expressed as a percentage (prev), and the 95% confidence interval for the prevalence as determined by the exact (also known as beta) approximation interval method, as implemented by the Python statsmodels module (v.0.14.4), also expressed as a percentage (CI). \*District Average is the equally weighted average of the districts for the region

Antifolate resistance mutations also saw small spatial and temporal shifts between 2021 and 2023. Mutations in DHFR are associated with resistance to pyrimethamine and are acquired sequentially. The DHFR mutations 51**I**, 59**R**, and 108**N** were all near fixation in all districts and all years surveyed, while the recent 164**L** mutation, which confers high-level pyrimethamine resistance as well as resistance to dapsone, showed more variable patterns across districts and years, ranging widely in prevalence from completely absent in Biharamulo in 2021 to 38.1% in Karagwe in 2022. The 164**L** mutation increased in frequency in four of the Eastern sites near Lake Victoria (**Figure 2A**), while decreasing in frequency in three Western sites near Rwanda. Mutations in DHPS are associated with resistance to sulfadoxine, with high-level resistance conferred by three mutations (437**G**, 540**E**, and 581**G**). DHPS 437**G** and 540**E** were both present near fixation across all districts and years surveyed, while the DHPS 581**G** mutation showed patterns of prevalence (**Figure 2B**) that were similarly widely varying with DHFR-164**L**, with prevalences ranging from completely absent in Biharamulo in 2021, to 48.1% prevalence in Kyerwa in 2021. The DHPS 581**G** and DHFR 164**L** mutations appear to be statistically significantly correlated with each other (Spearman r² 0.666, P=0.00003). A full list of prevalences, coverages, and confidence intervals for known key drug resistance point mutations can be found in **Table S3**.

## DISCUSSION

Between 2021 and 2023, malaria molecular surveillance (MMS) in the Kagera region revealed a concerning but nuanced picture of emerging ART-R. The WHO-validated K13 mutation R561**H** remains the predominant marker of ART-R, with evidence of both persistence in the original hotspot along the Rwanda border and gradual expansion eastward. Across 2,826 successfully genotyped isolates from seven districts the average R561**H** prevalence rose from 5.5% in 2021 to 6.9% in 2023. The highest and most consistent burden occurred in Karagwe (22.8–25.8%) and Kyerwa (14.4–13.9%), confirming sustained transmission in border districts. Importantly, R561**H** appeared for the first time in Muleba (5.0%) and Bukoba (0.7%) in 2023, with Ngara showing a modest rise (1.4% → 3.0%), indicating eastward spread into new districts. While the overall prevalence remains relatively low, any geographic expansion of K13 mutations warrants close attention, and it is somewhat reassuring that prevalence did not escalate further and did not involve additional districts over the two-year period.

Other validated K13 mutations also demonstrated early signals of spatial diversification. A675**V**, initially restricted to Karagwe, emerged in Muleba and Kyerwa by 2023. V568**G** appeared sporadically in Bukoba and Muleba, P441**L** emerged in Kyerwa and Misenyi, and the Horn-of-Africa–associated R622**I** mutation was detected once in Misenyi. Candidate and non-validated polymorphisms remained rare but increased slightly in frequency and numbers in later years, especially in Karagwe and Ngara. Collectively, these patterns indicate a parasite population undergoing continued diversification under local selection pressures.

Markers associated with ACT partner-drug tolerance and antifolate resistance displayed more modest temporal shifts. The MDR1 **N**86Y wild type allele, linked to lumefantrine tolerance, was nearly fixed across all districts. Conversely, CRT K76**T** mutation associated with amodiaquine and chloroquine resistance declined from 5.9% (2021) to 2.4% (2023), driven largely by decreasing prevalence in Ngara (24.2% → 9.7%). Antifolate resistance remains entrenched: early DHFR mutations (N51**I**, C59**R**, S108**N**) approached fixation, while high-level resistance markers associated with sulfadoxine-pyrimethamine resistance, DHFR I164**L** and DHPS A581**G,** exhibited substantial spatial and temporal heterogeneity. DHFR I164**L** ranged from 0% in Biharamulo to 38.1% in Karagwe, while DHPS A581**G** peaked at 48.1% in Kyerwa. Their strong correlation (r² = 0.666, P = 0.00003) suggests co-selection of high-level SP resistance haplotypes, a critical concern in regions implementing intermittent preventive treatment in pregnancy (IPTp). A recent meta-analysis indicated that IPTp with SP is associated with improved birth outcomes even when DHPS K540**E** is >90% but not when DHPS A581**G** is >10%.(van Eijk et al., 2019)

A defining feature of resistance evolution in Kagera is its pronounced microheterogeneity. K13 R561**H** prevalence exceeded 25% in Karagwe but remained undetectable in Biharamulo and Missenyi during the same period. Similarly, DHFR I164**L** and DHPS A581**G** ranged from near zero in western districts to over 40% in eastern districts near Lake Victoria. These stark gradients underscore that resistance is shaped by highly localized factors rather than uniform regional pressures. Test positivity can be used as a proxy for transmission intensity, noting that this correlation is limited in the districts with the highest and lowest transmission intensity.(Okiring et al., 2021) Transmission intensity is one likely driver: although six of seven clinics were situated in moderate-to-high transmission districts (Petro et al.,2024) (average RDT test positivity ≥20%), high transmission did not appear to suppress increases in ART-R mutations, contrary to some theoretical expectations regarding within-host competition. Local drug use patterns, stock variability, IPTp coverage, and human mobility patterns likely contribute to the localized emergence and persistence of resistant haplotypes.

The implications for surveillance and policy are clear. Region-level averages obscure district-level hotspots that may serve as reservoirs for broader dissemination. District-specific monitoring is therefore essential for early identification of emerging mutations, evaluation of partner-drug vulnerability, and targeted interventions such as modified treatment policies or intensified vector control. Micro-scale data will improve predictive modeling of resistance trajectories and inform evaluation of future strategies, including drug rotation, multiple first-line therapies, or triple ACT regimens.

There is growing urgency for such vigilance. ART-R in Africa may facilitate the emergence of ACT partner-drug resistance. In Uganda, *ex vivo* susceptibility to lumefantrine, Africa’s dominant partner drug, declined between 2019 and 2024 (Okitwi et al., 2025), and returning travelers have shown treatment failures accompanied by decreased in vitro lumefantrine susceptibility.(van Schalkwyk et al., 2024; Oliveira et al., 2025) These findings, combined with the increasing detection of multiple ART-R mutations in Kagera, underscore the critical role of MMS in protecting ACT efficacy. Tanzania also routinely implements intermittent preventive treatment in pregnancy (IPTp) in regions of high and moderate transmission, including Kagera. The anti-folate sulfadoxine-pyrimethamine (SP) remains the key drug for IPTp. Understanding the dynamics of key markers for high-level SP resistance is therefore necessary (**Table S3**).

This study also has important limitations. Sampling was uneven across years, with four districts not surveyed in 2022, limiting the resolution of temporal trends. Genotyping success varied by locus and year, potentially biasing prevalence estimates. The absence of whole-genome sequencing data prevents distinguishing whether R561**H** observations represent clonal expansion or multiple introductions. Sampling was limited to symptomatic clinic attendees, which may not fully represent community-level parasite diversity. Finally, transmission intensity estimates relied on test positivity rather than entomological or incidence data, constraining the interpretation of ecological drivers of resistance.

Despite these limitations, the data reinforce that ART-R remains an emerging but manageable threat—if surveillance remains strong. The TZ2 haplotype of R561**H**, first identified in Kagera in 2021, has since been detected in other regions of Tanzania, highlighting the potential for wider dissemination. Ongoing sequencing from all regions of Mainland Tanzania within the MSMT program will help clarify whether Kagera continues to seed R561**H** elsewhere. National malaria control programs in affected areas must therefore strengthen surveillance systems to ensure rapid detection and mitigation. Molecular markers of resistance, particularly K13 polymorphisms, are already being incorporated into modeling studies exploring the impact of alternative treatment strategies. Continued, granular MMS will be indispensable for informing these policy decisions and preserving ACT effectiveness across the region.

## Data Availability

Sequencing data for 2021 are publicly available under BioProject PRJNA1090883. Sequencing datasets for 2022 and 2023 will be released upon completion of manuscript submission (pending). Custom analysis scripts, individual participant level data, and genotyping call files are accessible on GitHub at: https://github.com/bailey-lab/msmt_dr_longitudinal_21-23/tree/mai

## Codes and Data Availability

Sequencing data for 2021 are publicly available under BioProject PRJNA1090883. Sequencing datasets for 2022 and 2023 will be released upon completion of manuscript submission (pending). Custom analysis scripts, individual participant level data, and genotyping call files are accessible on GitHub at: https://github.com/bailey-lab/msmt_dr_longitudinal_21-23/tree/main

## Author Contributions

**Data Curation:** AS, SM, AAF, NWY, MDS, FF

**Formal Analysis:** AS, SM, AAF, NWY, JAB

**Funding Acquisition:** DI, JJJ, JAB

**Investigation:** AS, SM, AAF, NWY, JJJ, DSI, JAB

**Methodology:** AS, AAF, NWY, JAB

**Project Administration:** CIM, FF, JJJ, DSI, JAB

**Resources:** JJJ, DSI, JAB

**Software:** AS, JAB

**Supervision:** JJJ, DSI, JAB

**Validation:** AS, JAB

**Visualization:** AS, AAF, NWY, JAB

**Writing – Original Draft Preparation:** AS, NWY, AF, JJJ, DSI, JAB

**Writing – Review & Editing:** All authors

## Acknowledgements

We extend our sincere gratitude to the following project staff who were involved in the implementation of the field and laboratory activities for the MSMT project; Raymond Kitengeso, Muhidin Kassim, Athanas Mhina, August Nyaki, Juma Tupa, Ildephonce Mathias, Grace Kanyankole, Gerion Gaudin, Honest Munishi, Oswald Osca, Anael Derick Kimaro, Ezekiel Malecela, Tilaus Gustav, Ruth Boniphace, Ramadhan Moshi, Kusa Mchaina, Emmanuel Kessy, George Gesase, Tumaini Kamna, Grace Kanyankole, Oswald Osca, Richard Makono, Hussein Semboja, Gineson Nkya, Daniel Chale, Richard Malisa, Sawaya Msangi, Ally Idrisa, Francis Chambo, Sharifa Hassan, Salome Simba, Hatibu Athumani, Ambele Lyatinga, Ally Idrissa and Amina Ibrahim. We are also grateful to the finance, administrative and logistic support team at NIMR: Christopher Masaka, Millen Meena, Beatrice Mwampeta, Neema Manumbu, Halfan Mwanga, Arison Ekoni, Twalipo Mponzi, Pendael Nasary, Denis Byakuzana, Alfred Sezary, Emmanuel Mnzava, John Samwel, Daud Mjema, Seth Nguhu, Thomas Semdoe, Sadiki Yusuph, Alex Mwakibinga, Rodrick Ulomi and Andrea Kimboi. We wish to thank all the study participants and parents and/or guardians of minors for providing consent and taking part in the study. We received great support in the implementation of the MSMT project from the regional and district authorities (particularly the Regional Administrative Secretaries, the Regional Medical Officers, the Regional Malaria Focal Persons, the District Medical Officers, and the District Malaria Focal Persons), NIMR management, and officials from the National Malaria Control Program and the President’s Office, Regional Administration and Local Government.

The following reagent was obtained through BEI Resources, NIAID, NIH: Genomic DNA from *Plasmodium falciparum*, Strain 3D7, MRA-102G, contributed by Daniel J. Carucci. The following reagent was obtained through BEI Resources, NIAID, NIH: Genomic DNA from *Plasmodium falciparum*, Strain 7G8, MRA-152G, contributed by David Walliker. The following reagent was obtained through BEI Resources, NIAID, NIH: Genomic DNA from *Plasmodium falciparum*, Strain K1, MRA-159G, contributed by Dennis E. Kyle.

## SUPPLEMENTAL MATERIALS

### SUPPLEMENTAL FIGURES

**Supplemental Figure S1:**
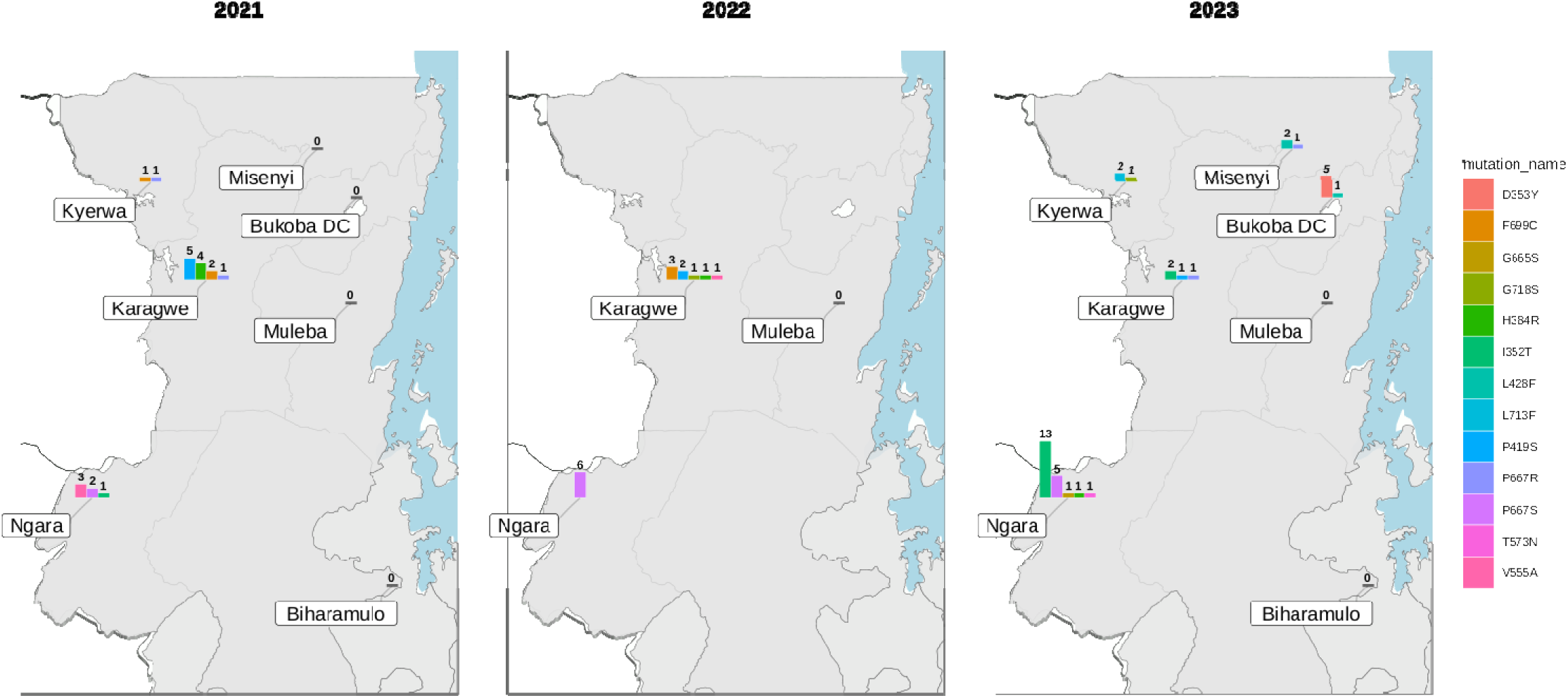
Noncanonical K13 Propeller Mutations. Each district shows a histogram with the number of samples containing each non-validated K13 mutation. The names of mutations associated with each bar in the histogram can be found by matching the color of a bar against the legend at the right.

### SUPPLEMENTAL TABLES

**To assess and view easily all supplementary tables compiled in one excel sheet.**

**Table S1**. The table shows validated or candidate artemisinin-resistance (ART-R) K13 gene mutation in Plasmodium falciparum parasites collected across various districts of the Kagera region (Tanzania) absent n 2021, 2022 and 2023. Values represent the number of mutant infections over total successfully genotyped samples, with 95% confidence intervals at district level.

**Table S2**: Non-Validated Non-Synonymous K13 Mutations Identified by Year and District (221-223). Values represent the number of mutant infections counts.

**Table S3:** District-Level Prevalence of Key Plasmodium falciparum Drug Resistance Mutations, 2021–2023. Values represent the number of mutant infections over total successfully genotyped samples, with 95% confidence intervals at district level.

